# Age-related loss of the Y chromosome is associated with sex hormone binding globulin and selected drivers of clonal hematopoiesis

**DOI:** 10.1101/2022.09.05.22279615

**Authors:** Ahmed A.Z. Dawoud, William J Tapper, Nicholas C.P. Cross

## Abstract

Mosaic loss of the Y-chromosome (LOY) is the most common somatic alteration in men. We aimed to assess the relationship between LOY and serum biomarkers in UK Biobank and explore the interaction with constitutional and somatic genetics. LOY was strongly associated with levels of sex hormone binding globulin (SHBG, β=0.12, P_FDR_= P = 7.44×10^−36^, adjusted for age, age squared, gender, smoking status, smoking intensity and principal genetic components), a key regulator of testosterone bioavailability associated with diverse disorders including cancer and autoimmune diseases. Furthermore, LOY was associated with total testosterone (TT, β=0.09, P_FDR_=2.23 × 10^−20^), but not bioavailable testosterone (P_FDR_=0.46) or free testosterone (P_FDR_=0.75). These relationships remained significant after sensitivity analysis that included comorbidities and body mass index (SHBG, β = 0.08, P_FDR_ = 4.61×10^−21^; TT, β = 0.05, P_FDR_ = 4.13 × 10^−9^). Mendelian randomisation suggested a causal effect of SHBG on LOY in BioBank Japan (P=6.58×10^−4^) but there was no evidence for an effect of LOY on SHBG (P=0.46). Assessment of cis-eQTLs for 13 genes associated with LOY identified two SNPs that were also associated with levels of SHBG, however only rs7141210 (imprinted *DLK1-MEG3* locus) modified the relationship between SHBG and LOY (rs7141210-T/T; P_interaction_=0.04) with low levels of SHBG seen specifically in men without LOY (β=-0.02, P=0.001), but not those with LOY (P=0.41). Age-related clonal hematopoiesis (CH) defined by somatic driver mutations was not associated with sex hormone levels but was associated with LOY at clonal fractions >30% (OR=1.52, *P*=2.92×10^−4^). *TET2, TP53*, and *CBL* mutations were enriched in high level LOY cases, but not *DNMT3A* or *ASXL1*. Our findings thus identify independent relationships between LOY, sex hormones and CH.

## Introduction

Age-related somatic loss of the Y-chromosome (LOY) in peripheral blood leukocytes is the most prevalent chromosomal alteration in men.^1^ LOY has been identified in as many as 20% of UKB male participants (median age = 58), but only 10% of these (2% of all men) had LOY involving >20% of leukocytes.^2-4^ A series of genome wide association studies (GWAS) have characterized inherited genetic variation that predisposes to LOY,^2,3,5^ with 156 independent loci explaining up to 34% of the heritability.^2,6^ LOY has also been linked to smoking behaviour,^1,3,7^ indicating that the environment as well as genetics is an important factor. LOY is associated with all-cause mortality, cancer mortality,^1,8,9^ and a wide range of non-malignant conditions.^1,10-13^ LOY is also associated with variation in blood cell counts ^4,14^ and has long been recognized as a recurrent clonal cytogenetic finding in hematological malignancies where, in the absence of other changes, it is associated with a good prognosis.^15^ LOY with large clone size has been linked to the presence of somatic mutations associated with hematological malignancies and an elevated risk of developing myeloid neoplasia in two recent, small studies of selected cases.^16,17^ It has been suggested that LOY might be a broad marker of genomic instability across different tissues and may exert its effects by altering immune cell function.^2,18^

Clonal hematopoiesis (CH) is a widespread phenomenon characterized by the presence of expanded clones of mutated blood cells,^19^ predominantly in individuals over the age of 60. Large autosomal chromosomal alterations including gains, losses and copy number neutral loss of heterozygosity (CN-LOH) inferred from SNP array data have been used to identify clonality,^20,21^ but CH is more commonly recognized by sequence analysis and the finding of pathogenic driver mutations associated with hematological malignancies, most commonly in the epigenetic regulators *DNMT3A, TET2*, and *ASXL1* but also a wide range of other genes.^22-28^ Broad screens by whole exome sequencing (WES) or whole genome sequencing (WGS) have revealed that clonality in the absence of known driver mutations (unknown driver CH) is even more prevalent than CH with driver mutations.^26^ Like LOY, CH has been linked to a wide range of malignant and non-malignant diseases.^23,24,29-33^ Most prominently, CH defined by autosomal chromosomal alterations or driver mutations is associated with a significant risk (hazard ratios of 10-12.9) of subsequent hematological malignancies,^20,21,24,25^ and recent studies showed a lineage-specific risk for mutations in genes associated with myeloid or lymphoid neoplasms.^34,35^ The clinical consequences of unknown driver CH have not been defined, and the reason for clonality in these cases remains unclear. Furthermore, the extent to which mosaic LOY can be considered as CH has not been defined. Although age is the major risk factor for both CH and LOY, it has become clear that there is significant overlap between genetic factors that predispose to both of these abnormalities. Broadly, inherited variants in cancer susceptibility genes and genes that are mutated in cancer feature prominently as risk factors for both CH and LOY.^2,3,6,36-38^ As for external factors, prior chemotherapy is associated with CH characterized by *PPM1D* and *TP53* mutations,^39^ whereas smoking is associated with *ASXL1* mutations as well as LOY.^40^

It is widely accepted that biochemical profiles change with age in a manner independent of specific disease states, for example ageing is associated with depletion of sex hormones.^41^ Sex hormone binding globulin (SHBG) is a glycoprotein that binds to steroids with high affinity, with both 5α-dihydrotestosterone and testosterone binding much more strongly than oestradiol.^42^ In men, circulating testosterone levels are regulated by SHBG, with on average 58% bound to SHBG, 40% bound to albumin, and 2% as free testosterone (FT).^43,44^ Binding to albumin is weak and so all non-SHBG-bound testosterone is considered as bioavailable testosterone (BAT).^45^ Ageing is associated with a decline in total testosterone (TT), FT and BAT and an increase in SHBG^41^ but the link, if any, between sex hormones and somatic genetic changes remains unexplored. In this study we aimed to determine if sex hormones and other serum markers are associated with LOY and to understand the interaction between biochemical factors with both constitutional and somatic genetics.

## Material and methods

### Study cohort

UK Biobank (UKB) is a large prospective cohort described in detail elsewhere^46^ involving approximately 500,000 individuals aged between 40 and 69 years at recruitment. Genome wide SNP data derived from peripheral blood leucocytes was available for most participants, and WES data for 200,631 participants at the time of analysis.

### Mosaic loss of the Y chromosome

A previous study used SNP data to identify males with mosaic LOY (n=44,588; 20% of evaluable males) using a method which compared allelic intensities for statistically phased haplotypes of the pseudo-autosomal region 1 (PAR1).^2^ This method for detecting LOY was considered to be less error prone than those based on the median genotyping intensity over the non-pseudoautosomal region of the Y chromosome and was able to detect mosaicism with a clonal fraction down to 1%.^2,47^ The spectrum of LOY was categorised according to clonal fraction by considering the median change of B-allele frequency (BAF) in PAR1, specifically BAFs of 0.026, 0.056, and 0.088 were previously shown to correspond with clonal fractions of 10%, 20%, and 30% respectively.^2^

### Biochemistry markers and sex hormones

Measurements of 29 biochemistry markers were available from serum samples collected on recruitment to UKB.^46^ Mass action equations were used to calculate FT and BAT from measurements of SHBG, TT and albumin as described.^48^ Further details regarding the biochemical assay methods and external quality assurance are available at https://biobank.ndph.ox.ac.uk/showcase/showcase/docs/serum_biochemistry.pdf.

### The relationship between biochemistry markers and LOY

We focused on the subset of male UKB participants who were evaluable for LOY assessment (n=222,835).^2^ The relationship between LOY and each of 31 biomarkers (29 measured and 2 calculated) was tested using multivariable linear regression in R. Continuous measures for each sex hormone were transformed into a normal distribution using inverse normal rank transformation and used as the dependant variable. For the basic model, the independent variables were LOY as a binary predictor, age, age squared, smoking status (never, previous, current), smoking intensity defined by average number of pack-years smoked by an individual over their adult lifetime ^49^ (Data-Field 20162) and the first 10 genetic principal components (10 PCA). For sensitivity analysis, we added more variables to the basic model, specifically binary variables derived from first occurrences of primary hypertension (ICD10:I10), insulin-dependent diabetes (E10), and non-insulin-dependent diabetes (E11), plus body mass index (BMI) as a continuous variable. Effect sizes were reported as beta coefficients (β) with 95% confidence intervals (95% CI). P-values were adjusted for 31 tests using the False Discovery Rate (FDR) method.^50^ The average measure of serum biomarkers in participants with LOY were compared to those in LOY free controls using Mann–Whitney U tests.

### Allelic SHBG score in UKB

Thirteen genetic variants were associated with circulating SHBG levels by a previous two-stage GWAS, including 10 variants that achieved genome wide significance plus 3 independent cis variants that were identified by conditional analysis of the *SHBG* gene.^51^ After excluding one SNP with heterogeneity towards females (*P* = 0.02, rs440837) and a second SNP on the X-chromosome (rs1573036), 11 SNPs were considered in relation to LOY. To consider both precision and power, two allelic scores were constructed. The first consisted of all 11 SNPs (rs12150660, rs1641537, rs1625895, rs6258, rs17496332, rs2411984, rs293428, rs780093, rs7910927, rs8023580 and rs4149056) while the second was based only on the 4 SNPs located in the vicinity of *SHBG* (rs12150660, rs1641537, rs1625895 and rs6258). Each score was calculated as the sum of number of risk factor-increasing alleles per SNP weighted by their corresponding genetic effect size. The allelic scores were calculated using Plink V1.9,^52^ and transformed into a normal distribution using inverse normal rank transformation. A multivariable logistic regression model adjusted for age, sex, smoking status and 10 PCA was used to assess the relationship between LOY status (binary, dependent) and *SHBG* allelic scores (independent, continuous).

### Mendelian Randomisation

Mendelian randomisation (MR) was used to assess the possibility of a causal relationship between SHBG and LOY using germline SNPs associated with circulating SHBG as instrumental variables, following the STROBE guidelines.^53^ Eight SNPs were used as instrumental variables without considering estimated genetic effect sizes for LOY in 95,380 men recruited to BioBank Japan (BBJ).^54^ Three of the eleven SNPs that were used to generate the allelic score (rs12150660, rs6258 and rs2411984) were excluded as genotypes were unavailable and/or non-informative in BBJ. We applied an inverse variance weighted model (IVW) with fixed effect using the TwoSamplesMR package.^55^ To explore the effect of SNP selection, we repeated the MR analysis using a genome wide significance threshold (*P* < 5×10^−8^) to select four SNPs (rs1641537, rs1625895, rs293428 and rs7910927) associated with SHBG.^51^ We also performed leave-one-out analysis to evaluate the effect of each SNP on the analysis. To assess the effect of LOY on SHBG levels we examined SNPs (n=50) previously associated with LOY ^54^ in BBJ. 10 SNPs were excluded as genotypes were unavailable and/or non-informative in UKB. The remaining 40 SNPs were used as instrumental variables without considering estimated genetic effect sizes for SHBG in UKB men.

### Identification of CH

A propensity score matching method in R ^56^ was used to select one control per case and to match for age in comparison to participants with LOY. Somatic mutations were called in LOY samples and matched controls using GATK (Version 4.1.9) and Mutect2 ^57^ to process individual CRAM files in the tumour-only mode. Following best practice guidelines (https://gatk.broadinstitute.org/hc/en-us/articles/360035531132), a Panel Of Normal (PON; version 23^rd^ August 2017) from the Broad Institute that were generated using Mutect2 on samples from the 1000 genomes project to identify recurrent artifacts, and germline variants from gnomAD were used to remove artefacts and germline variants. To identify putative somatic driver mutations, the analysis was restricted to rare variants with a minor allele frequency (MAF) <0.01 in gnomAD and a minimum number of reads supporting the mutated allele: 3 reads for point mutations and 6 reads for indels. Other methodological details for calling somatic variants are described in detail in our previous publication.^31^ Variants that satisfied either of the following two criteria were selected: first; recurrent driver mutations as defined in our previous study;^58^ second singleton variants that passed all Mutect2 filters with variant allele frequencies (VAF) between 0.1 and 0.2.^26^ Driver mutations were classified into myeloid or lymphoid according to a published list of genes associated with myeloid neoplasms (n=76) ^23^ and genes associated with acute lymphoblastic leukemia or chronic lymphocytic leukemia in the Cancer Gene Census (CGC),^59^ respectively (Table S1). Participants with variants in myeloid genes were considered as having myeloid CH (n=2,890), and those with variants in lymphoid genes as having lymphoid CH (n=532). Cases with mutations in genes involved in both myeloid and lymphoid neoplasms were considered as myeloid CH. To identify participants with evidence of clonality in the absence of pathogenic mutations in known driver genes, we also identified variants in all coding genes that were not defined as myeloid or lymphoid. Participants with singleton variants in any gene with VAF range between 0.1 and 0.2 not defined as myeloid or lymphoid were considered as having unknown driver CH (n=15,874).

### The relationship between LOY and CH

The association between LOY and all variants, driver variants, myeloid CH, lymphoid CH, and unknown driver CH were tested using logistic regression in R. We further assessed the relationship with LOY clone size categories (≤10%, 10%-20%, 20%-30%, and >30%) using Fisher’s exact tests. The strength of the association was reported as odds ratio (OR) with 95% CI. P-values were adjusted for 20 tests using the FDR method. To test the relationship at the individual driver gene level, the analysis was focused on genes mutated in ≥3 males with LOY in >30% of cells.

### Assessment of the co-existence of LOY and CH

The co-existence of CH and LOY was tested by assessing the relationship between the BAF for LOY and VAF of driver mutations. For cases with two or more mutations the highest VAF was used. To avoid excess CH with VAF between 0.1-0.2, the analysis for myeloid and lymphoid CH was restricted to VAFs detected for recurrent driver mutations as defined in our previous study.^31^ We assessed the relationship with LOY in ≤10% and >10% of cells. The dependent variables were VAF, age and smoking status (never, previous, current). The strength of the association was reported as β coefficient with 95% CI. P-values were adjusted for 6 tests using the FDR method.

### The relationship between CH and sex hormones

The association between CH and sex-hormone levels were tested using linear regression in R, with normally transformed sex hormone as the independent variable. The dependent variables were driver mutation state as a binary predictor, age, smoking status (never, previous, current), and 10 PCA. The association was reported as β coefficient with 95% CI. P-values were adjusted for 4 tests using the FDR method.

### Expression quantitative trait analyses

The eQTLGen database incorporates expression quantitative trait locus (eQTL) data from blood samples from a total of 31,684 individuals.^60^ To select a genetic proxy for gene expression, we filtered cis-eQTLs within a distance <1 Mb, and FDR < 0.05 and selected the SNP with the smallest FDR value, with no other genes showing a stronger association with the selected SNP to minimize horizontal pleiotropy. We restricted the analysis to directly genotyped SNPs with MAF >0.05 in UKB. 19 SNPs associated with LOY were associated with 27 genes by position, biological function, expression, or nonsynonymous variants in the gene.^3^ Our analysis was restricted to 13 of these genes for which an expression proxy was identified.

### Assessment of the interaction between eQTL, LOY and SHBG levels

The most significant eQTL SNP for each gene was encoded according to an additive model for the risk allele (0/1/2) in UKB and was factorized in each model with 0 as the reference (0/1 for heterozygous, and 0/2 if homozygous for the risk allele). The following statistical tests were applied, and adjusted for age, smoking, 10 PCA, and multiple tests by the FDR method. First, the relationship between each eQTL and LOY was assessed using logistic regression in R where LOY (binary) was the dependent variable and eQTL was the independent variable. Second, the relationship between eQTLs and SHBG levels was assessed using linear regression in R where SHBG (continuous) was taken as the dependent variable and transformed to a normal distribution using rank transformation, and eQTLs were the independent variable. Finally, if an eQTL was significantly associated with both LOY and SHBG, the interaction effect of the eQTL and LOY on SHBG regression was assessed by linear regression in R. Inverse normal rank transformed SHBG was considered as a continuous dependent variable, and each of eQTL, LOY, and eQTL x LOY (interaction effect) as the independent variable. To visualize the interaction effect in our models, we used ‘*interactions’* in R, a tool that was developed to interpret statistical interactions in regression models.^61^

## Results

### The relationship between LOY and biochemistry markers

To investigate the relationship between LOY and serum biomarkers, we used previously published calls of LOY that were generated by utilizing long-range phasing information to analyse allele-specific genotyping intensities of 1,239 variants in the pseudo-autosomal region 1.^2^ We restricted our analysis to the 222,835 males who passed QC, of whom 44,558 (20%) had LOY. Of these, the majority (n=31,952; 72%), had an estimated LOY clonal fraction of <10%. We compared the presence or absence of LOY with 29 biochemistry parameters that were directly measured by UKB, as well as estimated levels of FT (median = 0.21 nmol/L; range = 0.003 - 1.93) and BAT (median = 5.1 nmol/L; range = 0.09 - 45.68) that were derived from measurements of SHBG, TT and albumin.^62^

Univariate comparisons revealed that participants with LOY had higher median levels of alkaline phosphatase, apolipoprotein A, C-reactive protein, cystatin C, glucose, glycated hemoglobin (HbA1c), HDL cholesterol, TT, urea, SHBG and vitamin D. Lower medians levels in participants with LOY were seen for alanine aminotransferase, albumin, apolipoprotein B, aspartate aminotransferase, calcium, cholesterol, gamma glutamyltransferase, IGF-1, LDL direct, total bilirubin, total protein, triglycerides, urate and FT. (Table S2).

On initial multivariate analysis adjusted for age, age squared, sex, 10 PCA, smoking status, smoking intensity and FDR corrected for multiple testing, we found that LOY as a binary predictor was most strongly associated with elevated levels of SHBG (β = 0.12, 95% CI: 0.11 - 0.13, P = 7.44×10^−36^). SHBG binds steroids^42^ and it is notable that the second strongest positive association was with TT (β = 0.09, 95% CI: 0.07 - 0.11, P = 2.23 × 10^−20^). There was no association, however, between LOY and FT (P = 0.46) or BAT (P = 0.75) (Figure 1A and Table S3). Participants with LOY had higher levels of SHBG and TT (SHBG: median nmol/L = 41.54 vs 35.86, P < 0.001; TT: median nmol/L = 11.74 vs 11.58, P < 0.001; Mann-Whitney U tests) but lower levels of FT (median nmol/L = 0.19 vs 0.20, P < 0.001), and BAT (median nmol/L = 4.78 vs 5.18, P < 0.001). On sensitivity analysis corrected for hypertension, insulin dependent diabetes, non-insulin dependent diabetes, and BMI, LOY retained the positive association with SHBG (β = 0.08, 95% CI: 0.07 – 0.10, P = 4.61×10^−21^) and TT (β = 0.05, 95% CI: 0.04 – 0.07, P = 4.13 × 10^−9^; Figure 1B, and Table S3).Our observational results points to a direct relationship between levels of SHBG and LOY that cannot be explained by age, smoking, population stratification, common morbidities, BMI or free/bioavailable testosterone (Figure 2).

**Figure 1:**
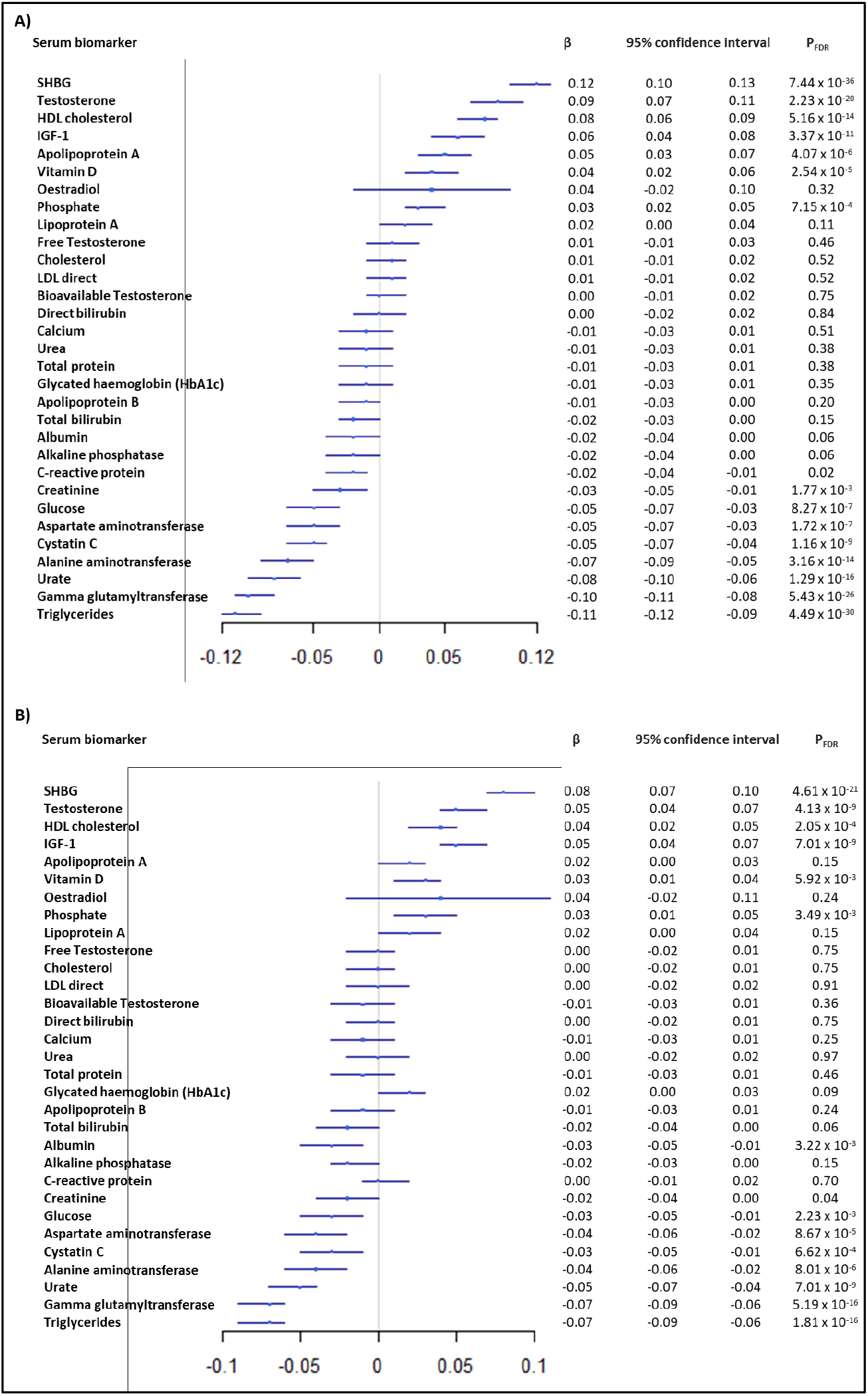
The relationship between LOY and biochemistry markers. The relationship between LOY and each of 31 biomarkers (29 measured and 2 calculated) was tested using multivariable linear regression in R in 222,835 UKB males. A) basic models: each linear model considered age, age squared, smoking status, smoking intensity 10PCA and multiple testing; B) sensitivity model: hypertension, insulin dependent diabetes, non-insulin dependent diabetes, and BMI were added to each basic model.

**Figure 2.**
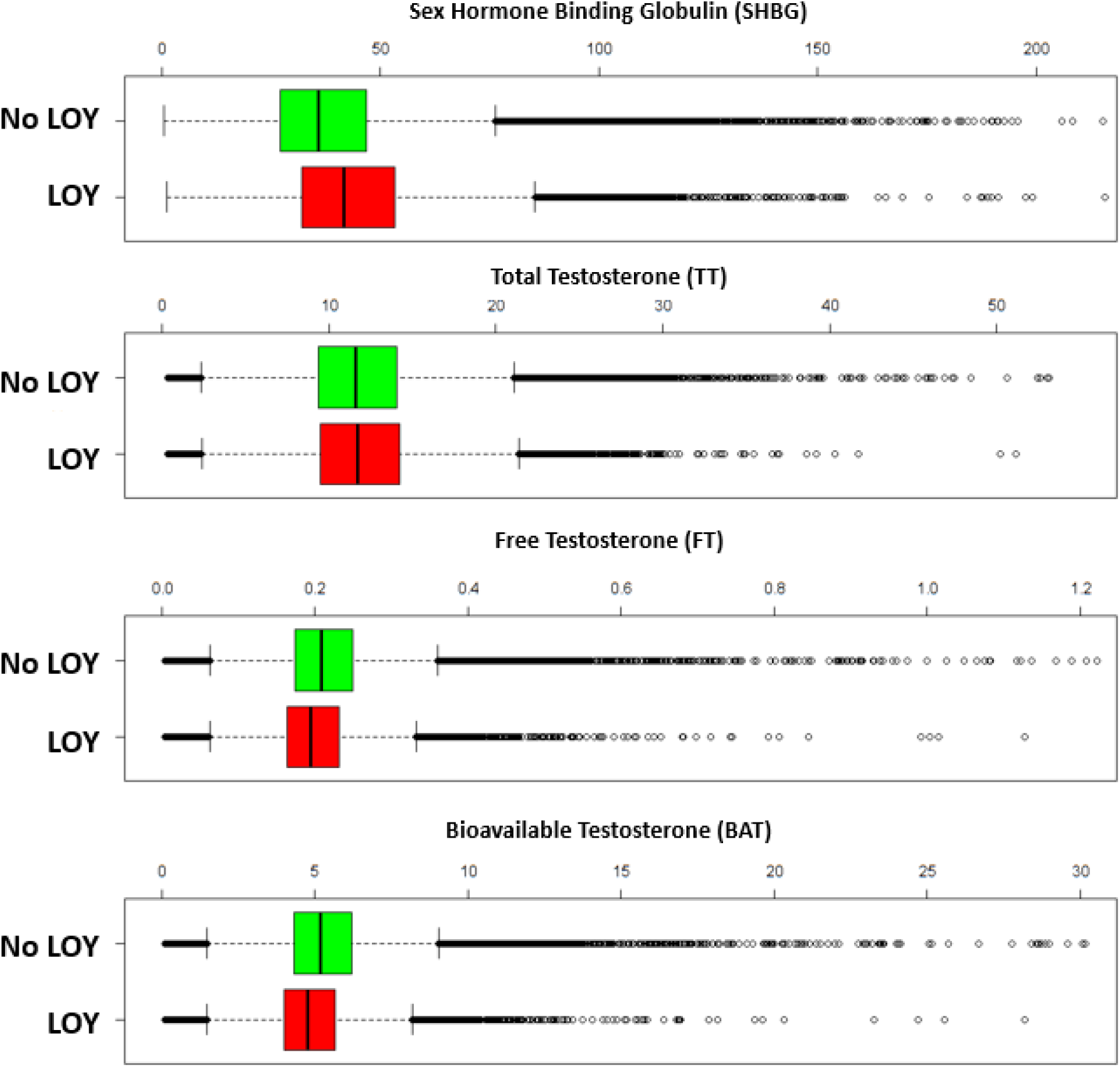
The relationship between LOY and levels of sex hormones. The box plots summarize serum sex hormone measurements in participants without LOY (n=178,277) and with LOY (n=4,458). SHBG: median nmol/L = 41.54 vs 35.86, P < 0.001; TT: median nmol/L = 11.74 vs 11.58, P < 0.001; FT: median nmol/L = 0.19 vs 0.20, P < 0.001; BAT: median nmol/L = 4.78 vs 5.18, P < 0.001.

### The relationship between genetically defined SHBG and LOY

Published GWAS have identified multiple genetic determinants of SHBG levels in serum.^51^ To understand the relationship between LOY and SHBG we generated allelic scores to summarize the genetic variation associated with SHBG and evaluated the scores as predictors of LOY. Since the allelic scores were derived from independent cohorts they represent unbiased instruments to assess the relationship with LOY in UKB.^63^ We found that genetically predicted SHBG was significantly associated with the finding of LOY in UKB using the score estimated from 11 independent SNPs (OR = 1.02, 95% CI: 1.01 – 1.04, P = 5.59×10^−5^).

To assess the possibility of a causal relationship between SHBG and LOY we performed MR analysis (Figure 3). In a liberal analysis, 8 independent SNPs were used to estimate the effect of SHBG on LOY (Table S4). Using an IVW model with fixed effect we identified a positive causal relationship (β = 0.15, 95% CI: 0.06 - 0.23, P = 6.58×10^−4^). In a more conservative analysis restricted to 4 SNPs associated with SHBG with P < 5×10^−8^, the effect of SHBG on LOY was confirmed (β = 0.17, 95% CI: 0.07 - 0.26, P = 7.28×10^−4^). Leave-one-out analysis (Table S5) found that significance was lost when rs7910927 at 10q21.3 within *JMJD1C* was excluded (liberal analysis: β = 0.08, 95% CI: -0.02 - 0.17, P = 0.13; conservative analysis: β = 0.07, 95% CI: -0.04 - 0.19, P = 0.22). To assess the possibility of bidirectional effect we utilized 40 SNPs with P < 5×10^−8^ associated with LOY in Japanese men ^54^ to measure their effect on SHBG levels in UKB men ^64^ but no significant relationship was found (β = 0.02, 95% CI: -0.88 - 0.13, P = 0.46).

**Figure 3.**
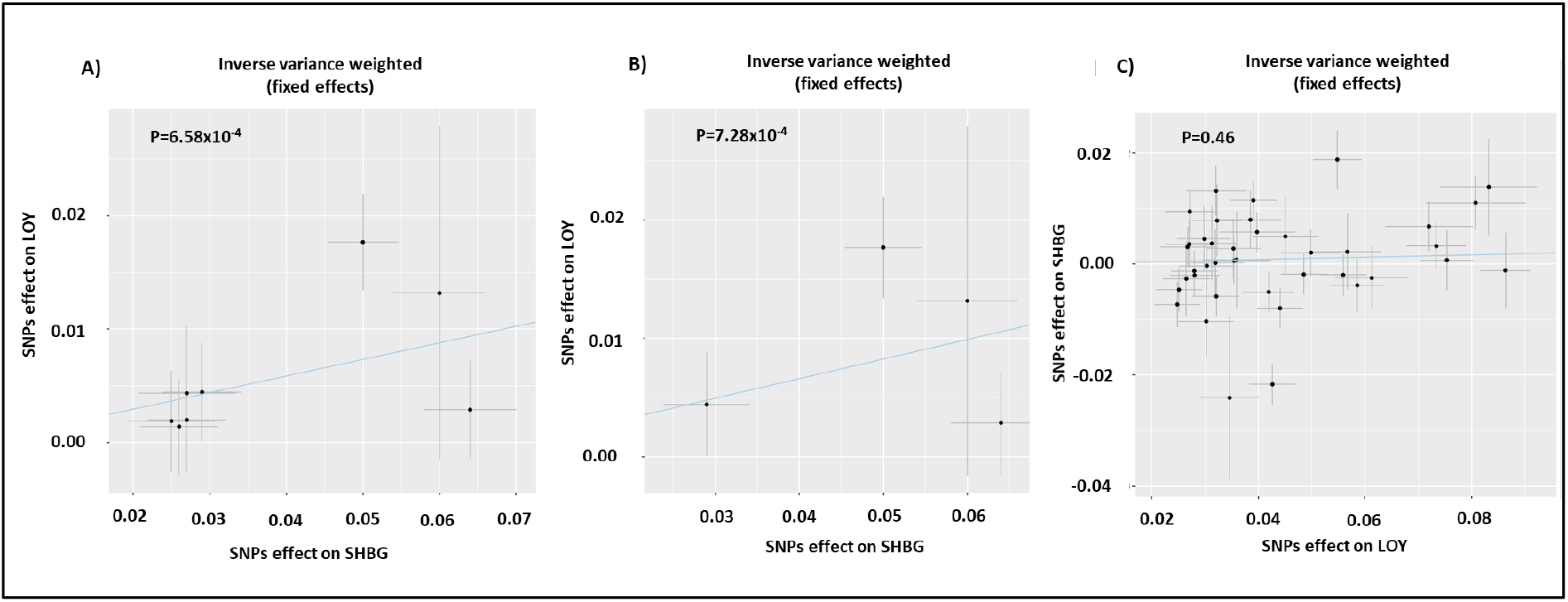
Mendelian randomisation using an inverse variance weighted model to estimate the causal relationship between SHBG and LOY. (A) Liberal analysis using 8 independent SNPs associated with SHBG. The IVW test estimated a significant positive effect of SHBG on LOY (P = 6.58×10^−4^). (B) Conservative analysis using 4 SNPs (P = 7.28×10^−4^). (C) analysis using 40 independent SNPs associated with LOY. The IVW test estimated no effect of LOY on SHBG (P = 0.46). The line of regression is indicated in blue, and the axes show β coefficients for SNP effects on SHBG and LOY.

### The effect of gene expression on the relationship between SHBG and LOY

To shed light on the possible mechanisms by which SHBG levels might promote LOY, we focused on 19 genomic regions robustly associated with LOY which had been confirmed in an independent GWAS.^2,3^. Using the eQTLGene ^60^ database we identified eQTL SNPs to serve as valid proxies for 13 of the 19 regions (*ACAT1, BCL2, DLK1, HM13, MAD1L1, QKI, RBPMS, SEMA4A, SENP7, SENP8, SETBP1, SMPD2, TCL1A*, and *TSC22D2*). We found that the heterozygous state of 8/13 eQTLs and the homozygous state of 9/13 eQTLs were associated with LOY in UKB. The eQTL for *TCL1A* at 14q32.13 had the strongest association with LOY status (rs11849538_G/C, OR = 0.83, 95% CI = 0.80 - 0.85, P = 6.32×10^−34^; rs11849538_G/G, OR = 0.63, 95% CI: 0.56 - 0.69, P = 1.48×10^−16^), as shown in Figure 4 and Table S6. Only 2 of the 13 genes, however, were significantly associated with levels of SHBG. *MAD1L1* at 7p22.3 was positively associated with SHBG (rs10247428_A/A, β = 0.02, 95% CI = 0.01 - 0.03, P = 0.003), but this was in the opposite direction to its relationship with LOY. The *DLK1-MEG3* eQTL at 14q32.2 was negatively associated with SHBG (rs7141210-T/T, β = -0.02, 95% CI = -0.03 - 0.007, P = 0.02) and also negatively associated with LOY (Table S6). Of these two signals, only rs7141210-T/T modified the relationship between LOY and SHBG (P_interaction_ = 0.04) (Figure 5, Table S7). There was no influence of rs7141210-T on TT, FT or BAT (Table S8) indicating that the interaction was specific for SHBG.

**Figure 4.**
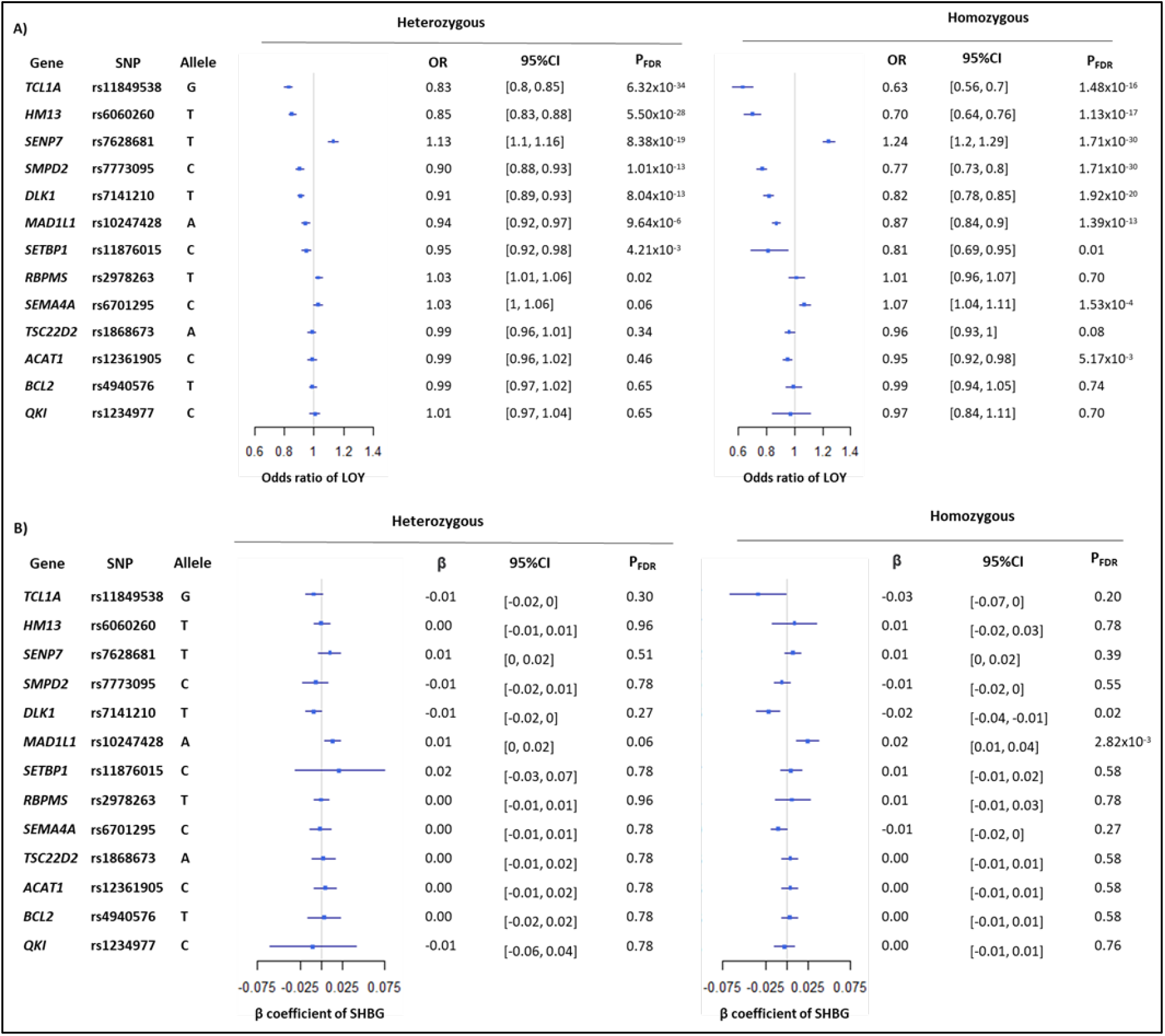
The relationship between the predicted expression of 13 genes and each of LOY and SHBG. eQTL SNPs were used as proxies for gene expression and as assessed as predictors for LOY (panel A) and SHBG levels (panel B) incorporating age, smoking status, and 10 PCA as covariates.

**Figure 5.**
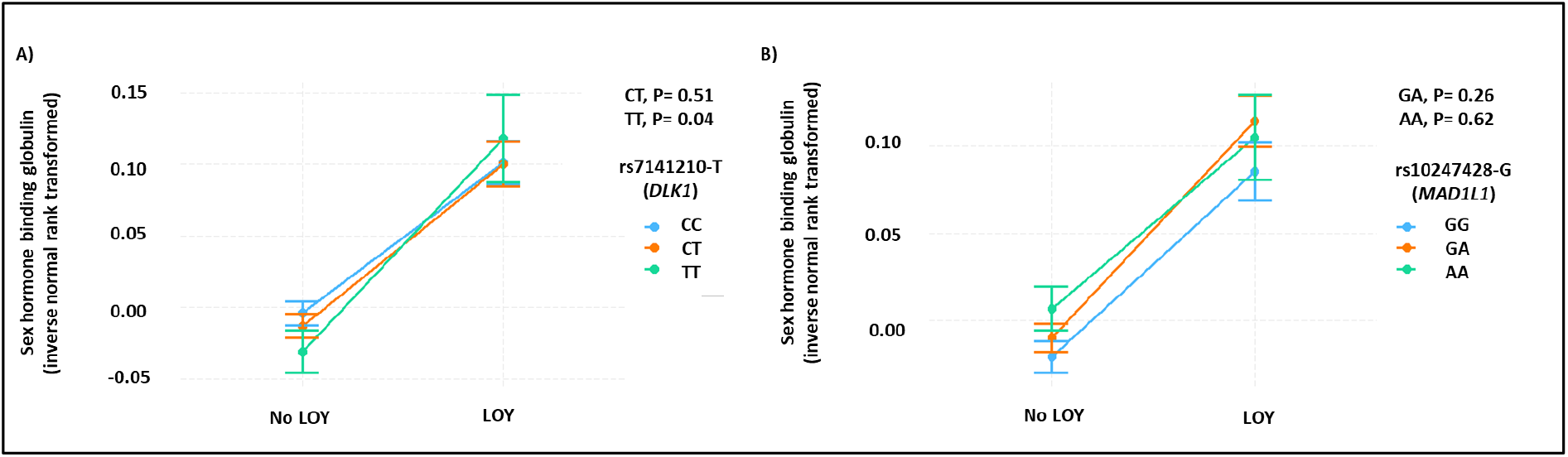
The interaction between genetically predicted expression of *DLK1*-*MEG3* and *MAD1L1*, according to LOY status and SHBG levels. Summary of SHBG values for men with and without LOY for (A) rs7141210 as a proxy for *DLK1-MEG3* expression and (B) rs10247428 as a proxy for *MAD1L1* expression. The interaction effect of each eQTL and LOY on SHBG was assessed by linear regression and was significant for rs7141210-T/T (green; left panel) in comparison to (C/C, blue). No significant difference was seen between rs10247428 genotypes.

### The relationship between LOY and CH

To assess the impact of somatic driver mutations and other markers of clonality on the relationship between SHBG, testosterone and LOY, we first assessed the relationship between somatic variants (driver and non-driver) and LOY. WES data was available for 17,759 participants with LOY, of whom 28% (n=4,981) were estimated to have an LOY clone size ≥ 10% of leucocytes. For comparison, we randomly selected UKB age-matched controls (n=17,702) that were negative for LOY. We identified recurrent somatic mutations in driver genes associated with myeloid CH and lymphoid CH, plus likely somatic variants in other genes that indicated clonality in the absence of known driver mutations, which we refer as unknown driver CH. Overall, the frequency of each CH subtype (myeloid, lymphoid, unknown) was similar between cases with LOY and controls. Striking differences emerged, however, when LOY was stratified by clone size. All CH (myeloid plus lymphoid plus unknown driver) was significantly associated with LOY in ≥ 10% of cells with a clear increase in the strength of the association with increasing LOY clone size (10-20% LOY, OR = 1.17, P = 1.81×10^−4^; 20-30% LOY, OR = 2.20, P = 4.25×10^−27^; ≥30% LOY, OR = 3.43, P = 2.42×10^−52^). Both myeloid CH (OR = 1.42, P = 4.52×10^−3^), and lymphoid CH (OR = 1.93, P = 0.01) were significantly associated with LOY in ≥30% of cells but not LOY of smaller clone size. The relationship was most prominent, however, for unknown driver CH (OR = 2.46, P = 5.09×10^−34^). Full results are presented in Figure 6 and Table S9. None of the three CH subtypes were associated with SHBG or the three measures of testosterone, indicating no effect of driver mutations on the relationship between LOY and sex hormones (Table S10).

**Figure 6.**
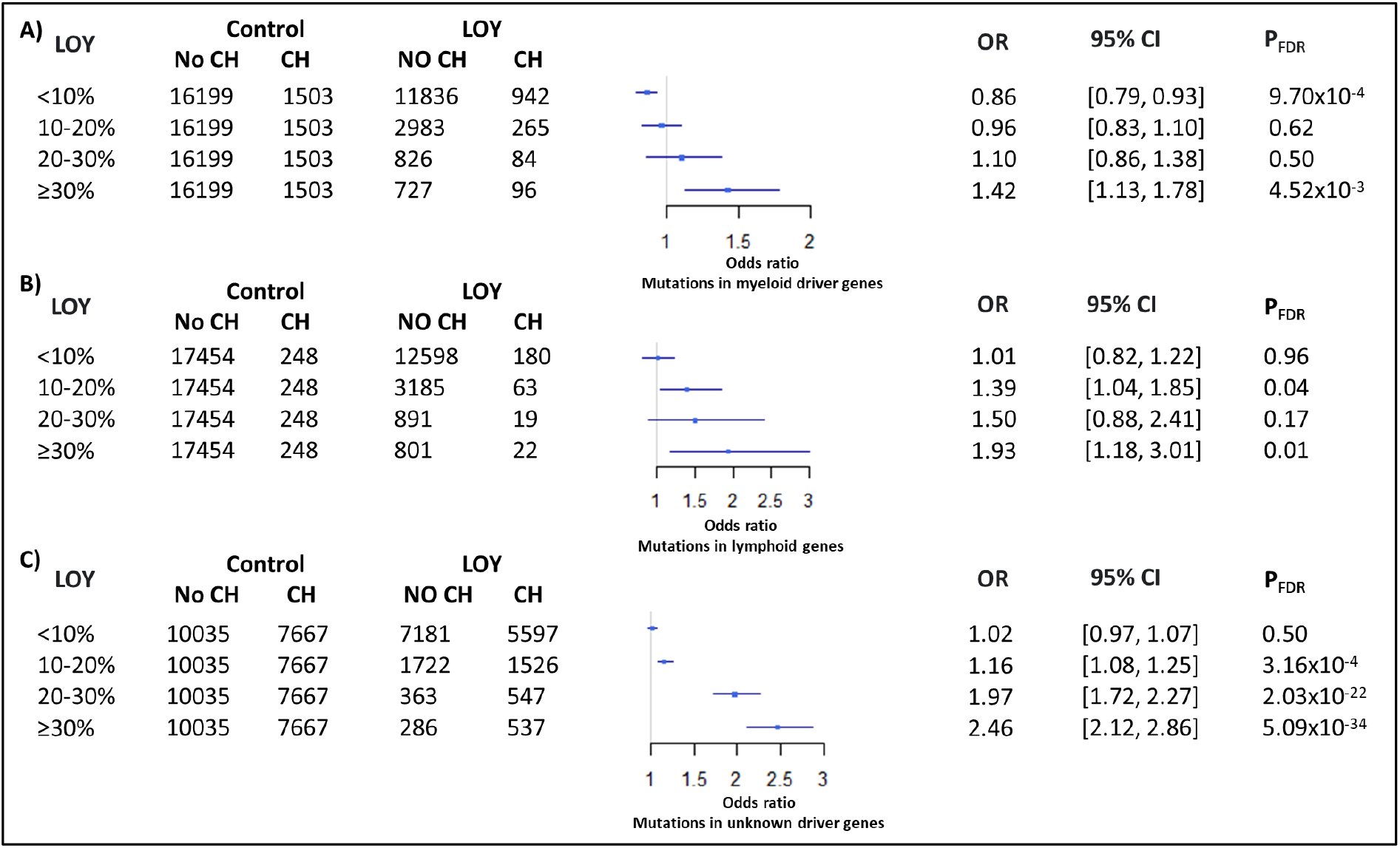
The relationship between LOY and CH. LOY was stratified according to the clonal size (<10%, 10%-20%, 20%-30%, and ≥30%) and the proportion of participants with CH within each group was compared with controls. (A) myeloid CH, (B) lymphoid CH, (C) unknown driver CH.

To understand the relationship between CH and LOY in more detail, we assessed the association between somatic mutations in specific driver genes in participants with LOY in >30% cells (n=823) compared to LOY free controls (n=17,702). *TET2* was the most significantly enriched mutated gene in LOY cases (4% versus 1.5% in controls, OR = 2.64, P = 9.58×10^−5^) with *TP53* (OR = 6.96, P = 7.62×10^−3^) and *CBL* (OR = 7.43, P = 0.04) mutations also showing a significant enrichment (Table S11). Other genes, including *DNMT3A* and *ASXL1*, showed no enrichment in high level LOY cases.

The possibility that LOY and CH as defined by somatic mutations might co-exist in the same clone was assessed by analysing the relationship between LOY BAF and the VAFs of driver mutations. Figure 7 shows a summary of the results at different ranges of LOY. Myeloid CH VAFs predicted BAF levels in samples with LOY >10% (β = 0.10, 95% CI = 0.05 - 0.15, P = 1.12×10^−4^). Similar results were seen for lymphoid CH (β = 0.20, 95% CI = 0.06 - 0.35, P = 0.02) and also unknown driver CH (β = 0.19, 95% CI = 0.14 - 0.24, P = 3.72×10^−12^), which by definition was restricted to VAFs of 0.1 - 0.2 (Table S12).

**Figure 7.**
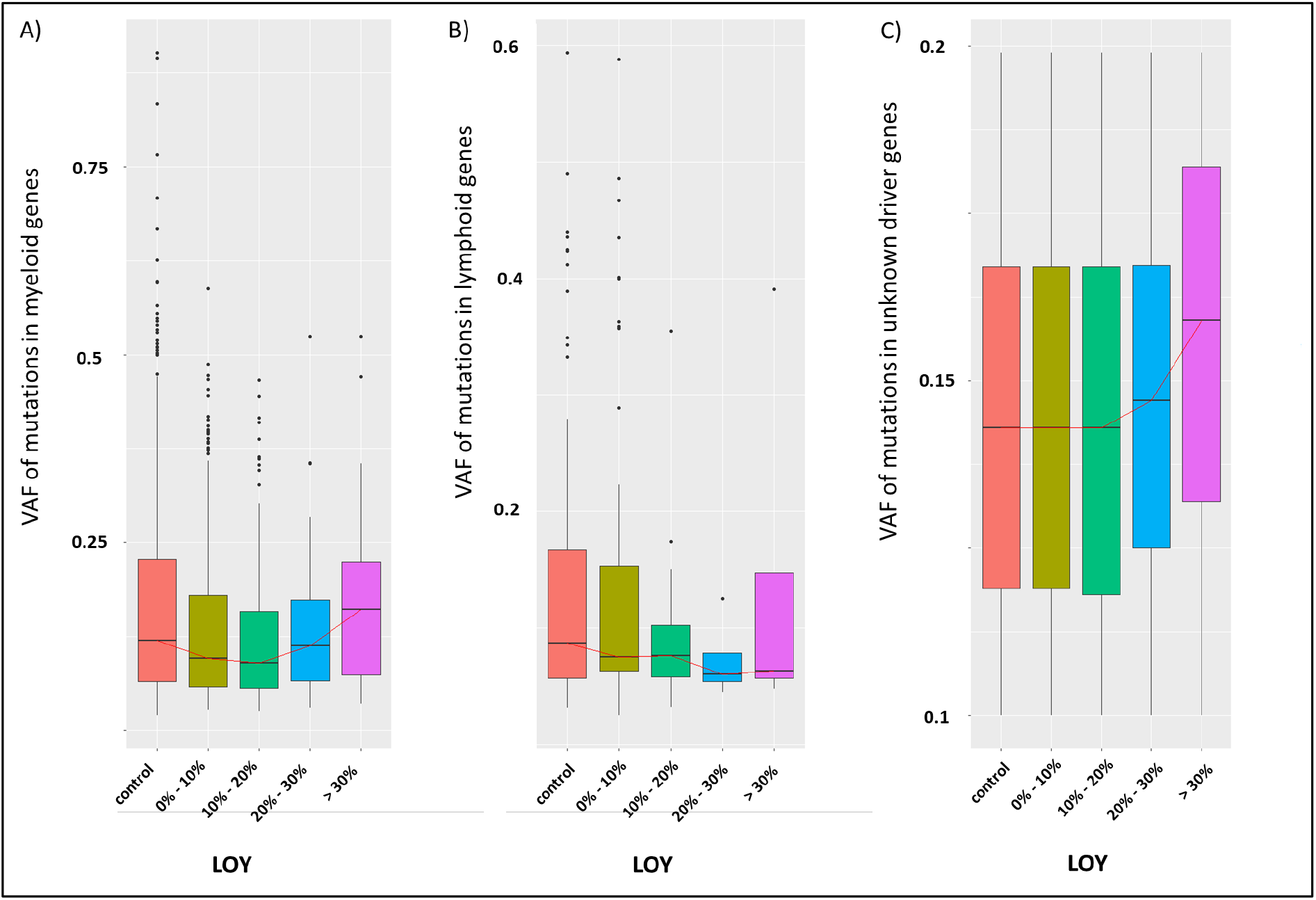
The relationship between LOY clonal size and CH VAFs. Boxplots summarizing the distribution of VAFs of somatic mutations in controls and cases with LOY broken down by clone size. (A) myeloid CH, (B) lymphoid CH, (C) unknown driver CH. The red lines connect median values.

## Discussion

Age-related mosaic LOY in peripheral blood leukocytes is known to be influenced by both genetic and environmental factors. We have found that LOY is also strongly with multiple serum biomarkers, most notably levels of SHBG, and that this association is independent of known confounders (age, age squared, smoking history, common comorbidities, BMI and the first 10 principal genetic components). Furthermore, we found genetic evidence to support the hypothesis that SHBG levels are causally linked to LOY, but no evidence that LOY has any effect on SHBG. SHBG regulates the level of circulating testosterone and, although we found that both FT and BAT were lower in men with LOY compared to those without LOY, there was no significant relationship between LOY and either FT or BAT on multivariate analysis. This finding is inconsistent with the free hormone hypothesis, which proposes that only the unbound fraction of testosterone is biologically active in target tissues,^65^ and instead suggests that involvement of other pathways such as binding and internalization of SHBG-bound testosterone by specific cell types, including B-cells.^66,67^ The mechanism by which SHBG promotes LOY is unclear, but from a genetic perspective the effect is not explained by variation at *SHBG* alone. Other loci are involved, particularly *JMJD1C* which encodes a histone demethylase previously linked to SHBG levels.^51^

To understand the influence of genetic factors on the relationship between SHBG and LOY in more detail, we focused on genetically predicted expression of genes linked to the development of LOY. We found that rs7141210-T, a marker for expression of genes in the *DLK1–MEG3* region at 14q32,^60^ was associated with elevated LOY and SHBG, and that homozygosity for the T-allele of rs7141210 modified the relationship between LOY and SHBG. *DLK1-MEG3* is a large and complex imprinted cluster of genes and non-coding RNAs. The methylated paternally derived chromosome expresses the protein-coding genes *DLK1, RTL1* and *DIO3*, while the non-methylated maternally derived chromosome expresses the non-coding genes *MEG3, MEG8, asRTL1*, multiple miRNAs and lncRNAs.^68^ Constitutional uniparental disomy (UPD) at 14q32 is associated with the developmental disorders Temple syndrome (maternal UPD) and Kagami–Ogata syndrome (paternal UPD) whereas somatically acquired paternal UPD is associated with CH and myeloid malignancies.^69^ Genome-wide significant signals have been identified near *DLK1* in association with CH defined by acquired 14q UPD ^70^ and somatic driver mutations ^71^ as well as LOY.^2,3,54^ Furthermore, a parent of origin specific effect of rs1555405-A linked to differential methylation has been defined in relation to platelet counts.^72^ This SNP is in linkage disequilibrium with rs7141210 (D’=1, R^2^=0.7), with rs7141210-T allele being correlated with rs1555405-A allele. Collectively these findings suggest the possibility of a parent of origin impact of rs7141210-T on the relationship between SHBG and LOY.

We have defined the relationship between CH and LOY in UKB. LOY in ≥30% of cells was associated with both myeloid and lymphoid CH, with 14% of affected individuals having one or more somatic driver mutation compared to 10% of controls (P = 2.92 × 10^−4^; Table S9). At the level of individual genes, the most striking finding was that mutated *TET2* was associated with LOY, but not *DNMT3A* or *ASXL1*. Collectively, mutations in one of these genes accounts for 90% of cases of CH defined by sequence analysis, and our findings are consistent with the notion that CH with *TET2* mutations is different from CH with *DNMT3A* or *ASXL1* mutations. With larger studies, specific disease associations are emerging, for example CH with *TET2* mutations has been linked to chronic obstructive pulmonary diseases,^73^ but not CH with *DNMT3A* mutations.

Most strikingly, however, unknown driver CH was seen in 65% of cases with high level (≥30% of cells) LOY compared to just 43% of controls (P = 5.09 × 10^−34^). Overall, 80% of cases with high level LOY had mutational evidence of clonality, with LOY in ≥10% of cells clearly associated with unknown driver CH. For the first time, therefore, our findings provide broad molecular confirmation that LOY ≥10% is indeed clonal, and we predict that comprehensive sequencing by WGS will confirm clonality in most cases. Importantly for our study, neither overall CH nor any CH subtype was associated with SHBG or measures of testosterone (Table S10). The driver of clonality in cases with LOY and unknown driver CH remains unclear but we found that the degree of LOY was strongly predicted by the VAF of the somatic variants used to define CH (P = 3.72×10^−12^). This suggests that LOY might itself be a driver of clonality, as has been postulated recently from whole genome sequence data of single cell-derived hematopoietic cells colonies,^74^ and that LOY therefore accounts for an appreciable proportion of unknown driver CH.

In summary, we conclude that (i) SHBG is associated with LOY, but this relationship cannot be explained by the free-hormone pathway; (ii) SHBG has a causal effect on LOY, but LOY has no effect on SHBG; (iii) expression of imprinted genes in the *DLK1-MEG3* locus alters the relationship between LOY and SHBG, but has no significant effect on other sex hormones; (iv) CH does not explain the relationship between LOY and SHBG, but confirms that LOY is clonal.

## Data Availability

All data produced in the present work are contained in the manuscript

## Supplemental data

Supplemental Data includes 12 tables

## Acknowledgments

AAZD was supported by a Lady Tata International Award. The authors acknowledge the use of the IRIDIS High Performance Computing Facility and associated support services at the University of Southampton.

## Author contributions

The study was designed by AAZD, WJT and NCPC. AAZD performed the data analysis in conjunction with WJT. All three authors refined the analysis and contributed to writing the paper.

## Web resources

Access to the UK Biobank Resource is available by application (http://www.ukbiobank.ac.uk/). eQTLGen database is available from (https://www.eqtlgen.org). PLINK (v1.9) is available from (https://www.cog-genomics.org/plink/1.9/). TwoSampleMR guide is available from (https://mrcieu.github.io/TwoSampleMR/). The code used for calculating FT and BAT is available from (https://labrtorian.com/tag/free-testosterone/). Further details regarding the biochemical assay methods and external quality assurance are available at https://biobank.ndph.ox.ac.uk/showcase/showcase/docs/serum_biochemistry.pdf.

